# Peritraumatic Distress of COVID-19 on Physicians in Bangladesh: Implications and Policy Recommendations

**DOI:** 10.1101/2021.07.05.21250138

**Authors:** Zafar Ahmad, Abdullah al Kium, Md Ripon Ahammed, Md Montaser Hamid, Sarmina Tarannum, Md. Ruhul Amin, Md. Daharul Islam

## Abstract

COVID-19 pandemic has been an ultimate test of resource management for any governance, especially in the healthcare system. Bangladesh, being a developing country and with very limited resources, is fighting the COVID-19 pandemic. The frontline workers, especially the physicians and nurses are going through immense physical and psychological stress during the pandemic. Social unawareness, the absence of strict preventive policies, increasing workload, and the lack of resource management are making the frontline healthcare workers extremely vulnerable to COVID-19. In this paper, we present the outcome of our study on peritraumatic distress of COVID-19 among physicians in Bangladesh. Based on the user study, we have identified a number of key factors behind the peritraumatic distress and psychological stress caused by COVID-19. Our study shows, more than 78% respondents are suffering from peritraumatic psychological distress. We also recommended some very important and yet easy to implement policies to reduce the peritraumatic stress of the physicians of Bangladesh. These policy recommendations were a result of the survey analysis and the suggestions from the COVID-19 designated physicians.

## Introduction

Like most of the countries in the world, all the major sectors of Bangladesh have been heavily impacted by the destruction caused by the Coronavirus Disease 2019 (COVID-19). Among them, the healthcare sector has taken the biggest hit from the damages imposed by this epidemic. Even before the pandemic, the existing healthcare system of Bangladesh was struggling in terms of manpower and skilled workers. In Bangladesh, there are only 3.05 physicians and 1.07 nurses per 10,000 population [12]. This sector also suffers from a lack of resources, reliability, empathy, and responsiveness [8], [9]. The crisis that emerged due to COVID-19 made this incompetent scenario of the healthcare sector of Bangladesh vividly clear [10]. The healthcare workers who remain under consistent pressure due to the inept working condition are now at even greater risk due to COVID-19. Being responsible for providing healthcare during this pandemic, they are in the frontline of facing the severity of COVID-19. Social unawareness, the absence of strict preventive policies, increasing workload, and inadequate testing kits and equipment are making the frontline healthcare workers extremely vulnerable to COVID-19 [11]. This vulnerability is resulting in an enormous amount of mental and physical stress for them.

Previously, it had been seen that working in hospitals during pandemics including the ongoing COVID-19 crisis can cause severe psychological distress and post-traumatic stress among healthcare workers [13]. Even before COVID-19, depression, anxiety, and a suicidal tendency among the physicians were highly prevalent compared to other professions [16], [17]. This is also true for medical students and residents [18], [19]. They are more susceptible to depression and burnout than most of the people who are in other career paths [18], [19]. The situation that emerged due to COVID-19 made the mental health aspects of these people even worse. High infection and death rate, poor understanding of the virus, and quarantine measures can classify COVID-19 as a traumatic event that is causing acute stress disorders among healthcare workers [14]. This may eventually develop into chronic post-traumatic stress disorder [14]. According to the latest findings, the prevalence rate of trauma-related stress among frontline healthcare workers due to COVID-19 is the highest and it ranges from 7.4% to 35% [15]. This high prevalence of stress can have prolonged impacts on the healthcare sector and the people associated with this sector.

In Bangladesh, the spread of COVID-19 started in March 2020 and increased very rapidly in the following four months [11]. Till 16 December 2020, about 495,841 people have been diagnosed with COVID-19 in Bangladesh [20]. The number of total deaths is 7,156 and so far 429,351 people have recovered from this disease [20]. However, the actual numbers of the pandemic are much worse than the available data. The testing rate (1.827%) in Bangladesh is one of the lowest in Asia [21]. Moreover, most of the patients (79%) are getting treatment from their homes due to the fear of maltreatment from hospitals [22]. As a result, it is not possible to get a clearer idea about the actual scenario of COVID-19 in Bangladesh.

This overwhelming scenario of the COVID-19 crisis is imposing unbearable physical and mental stress on the healthcare workers [20]. Most of the hospitals are unable to provide adequate protective materials, ventilators, and testing kits. Moreover, physicians and nurses are responsible to make vital ethical decisions while providing services to the patients during this pandemic [15]. This is even more prevalent in a resource-scarce country like Bangladesh. The percentage of physicians’ deaths among the COVID-19 fatality cases in Bangladesh was reported to be the highest in the whole world [6]. All these circumstances are creating enormous stress among the healthcare workers of Bangladesh.

In this paper, we present the outcome of our study on peritraumatic distress of COVID-19 among physicians in Bangladesh. Based on the user study, we have identified a number of key factors behind the peritraumatic distress and psychological stress caused by COVID-19. We conducted a survey following the questionnaires of previously published research workers in [3] and [4]. We received 241 responses in total from healthworkers among which 237 responses is from physicians and 4 responses are from nurses/hospital workers. Therefore, for the remaining text, we will mainly focus our analysis on the physicians’ responses. In this paper, we present the different statistical analyses on the physician’s response to our survey contributing towards an understanding of the real mental health scenario of healthcare workers. Based on the findings, we also recommend policies that will alleviate the severity of the COVID-19 imposed psychological stress among the healthcare workers.

### COVID-19 Crisis and Peritraumatic Distress Among Physicians in Bangladesh

Any disaster results in damage or distress among the affected sectors. For COVID-19, the most affected area is the healthcare sector. The first wave of the COVID-19 overwhelmed the healthcare sectors all over the world. However, for Bangladesh, the situation was even more challenging. Shortage of PPEs, testing kits, oxygen supplies, and ICU facilities resulted in huge distress to the general people including healthcare workers of Bangladesh. Thus for physicians, providing healthcare became the riskiest task during the pandemic.

To articulate the COVID-19 crisis in Bangladesh, we analyzed the transmission dynamics of the pandemic using SEIR model [1], [2]. **Figure 1(a)** shows the reproduction rate (*R*_*t*_), TPR, drop in mobility, and daily growth of COVID-19 patients over time. Though the *R*_*t*_ value is around 1, but the TPR is more than 10%. It is at least double compared to WHO’s recommended TPR of (5%). It clearly shows that the number of reported COVID-19 positive cases alone is not a good indicator of the pandemic’s extent in Bangladesh. We speculate that until the positive test rate doesn’t go below 5%, the number of positive reported cases is an underestimation of the actual number of infections. Therefore, we assume there are possibly too many patients with COVID-19 symptoms who are not diagnosed with the virus either due to the lack of testing facilities or other economic and societal issues for which people refrain themselves from seeking medical treatment. This situation puts our frontline workers in danger of getting exposed to the disease; therefore, adding anxiety, stigma, and psychological distress to our fellow Physicians.

**Figure 1.**
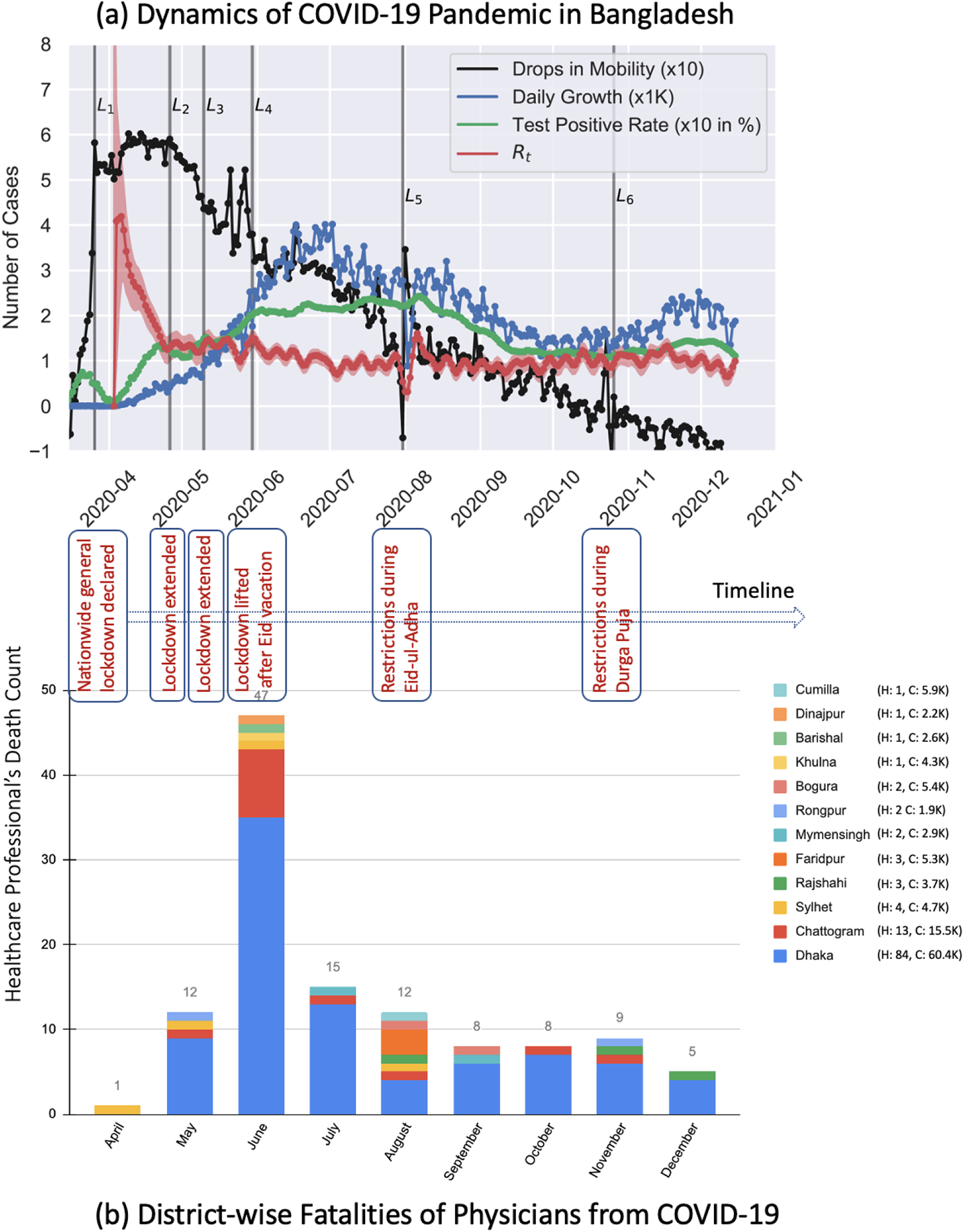
In Figure (a), we present the timeline of COVID-19 outbreak in Bangladesh and death toll of physicians during this time. It can be observed that even though mobility has increased (presented as the drops in Google mobility data) since the month of June, and the test positivity rate (TPR) is above 10%. Interestingly, the *R*_*t*_ is still around 1.0, which contradicts the higher TPR. In Figure (b), we present the distribution of physicians death in different districts of Bangladesh. In each of these districts there is at least one specialized hospital for COVID-19 treatment.

Bangladesh suffered from the first death of a physician, Dr. Moin Uddin Ahmed, on April 15th, 2020. Although he was an assistant professor in the medicine department of Sylhet MAG Osmani Medical College Hospital, unfortunately, he was the first known COVID-19 patient in the Sylhet division. He had to shift to the Dhaka division for an ICU facility and ventilated for three days in the Kurmitola General Hospital. His death left a devastating impact on the whole healthcare system as he might have contracted the disease while treating patients at the hospital without having enough protective equipment. Since then, according to the report of the Bangladesh Medical Association (BMA), 2,890 doctors got infected and 125 doctors lost their life to COVID-19 till January 7th, 2021 (**Figure 1(b)**). Thus the fear of catching COVID-19, extra-working hours, and physical and mental stress has become the source of the physicians’ peritraumatic distress.

Therefore, to raise the concerns about the mental health status of the healthcare workers in Bangladesh we have conducted the peritraumatic distress study on physicians. COVID-19 Peritraumatic Distress Index (CPDI) index score captures the stress index for the general mass population. But It is not sufficient to capture the unique physician-specific circumstances. We used the physician-specific questionnaire from the study of psychological stress of medical staffs during the outbreak of COVID-19 [4] and modified accordingly for the physicians of Bangladesh. A self-report questionnaire was designed to assess the CPDI among the healthcare workers during the first wave of the COVID-19 pandemic. Our study shows, more than 78% respondents are suffering from preritraumatic psychological distress. We also recommended some very important and yet easy to implement policies to reduce the peritraumatic stress of the physicians of Bangladesh. These policy recommendations were a result of the survey analysis and the suggestion from the COVID-19 designated physicians.

## Methodology

This study of COVID-19 peritraumatic distress on the physicians in Bangladesh is the first nationwide survey conducted to assess the COVID-19 frontline healthcare workers’ peritraumatic distress in Southeast Asia. It was well-received in the healthcare community and was featured in the most popular news portal for healthcare workers in Bangladesh [5]. We discuss our approaches for conducting the survey including analyzing the survey data below.

### Data Collection

A self-report questionnaire was designed to assess the CPDI among the healthcare workers during the first wave of the COVID-19 pandemic. We had created an internet-based data collection form following a similar study, [3]. Four physicians, co-authors of this manuscript, who were designated COVID-19 healthcare workers, were involved in updating the survey in the context of our study objectives. The survey questionnaires were modified accordingly to conduct the study for the healthcare workers of Bangladesh.

At the beginning of the survey, the objective of the study and question settings were explained, and then an informed consent of the study was taken from the participants. The criteria of selecting the participants were that only those adults who were directly involved to provide medical treatment physically to the COVID-19 patients in Bangladesh. To start filling out the questionnaire, participants were needed to choose the “agree” option; otherwise, the questionnaire could not be filled out. Confidentiality was managed by placing anonymous coding for each self-report questionnaire. Following the consent of survey participants, we collected the demographic information of each participant. Then, we asked users’ input for a set of peritraumatic distress questionnaires [3]. In addition, we incorporated medical staff specific psychological stress questionnaires following the study published in [4].

To conduct the survey, an Internet-based form was launched on the 23rd of June, 2020. We reached out to doctors who were actively working during the pandemic via social networks, academic emails, WhatsApp, and physicians news portal [5]. In the long haul of careful data collection, this study received a total of 237 valid responses from doctors who were actively working during the first-wave of COVID-19. The survey was conducted until August 31, 2020.

### Statistical Analysis

The online survey consisted of two main sections. The first section included social-demographic information, occupation and work history, and current physical status. The second section included three sets of peritraumatic distress questionnaires: concerns to be infected by COVID-19 from colleagues, patients, and visitors; physical uneasiness including changing food, sleeping, and conversing habit; and personal opinion about the improvement of the current situation.

The CPDI score was calculated by adding the responses of 24 CPDI questions from the survey dataset. In these questions, the physician responses were recorded on the Likert scale (0 to 4). Adding those 24 scores, we calculate the overall CPDI score of a physician. An overall CPDI score < 28 indicates mild distress. Whereas a score between 28 and 51 indicates moderate distress. Lastly, a person with a CPDI score ≥ 52 means the physician suffers from severe distress [3]. The validity of CPDI has been verified by researchers and the Cronbach’s alpha of CPDI is 0.95 (*p* < 0.001). In the frontline worker specific psychological stress questionnaire, there are nine questions to analyze in our dataset. The responses to those questions are also on the Likert scale (0 to 4). We analyzed these physician-specific psychological distress questionnaires using the T-test to find out any significant differences among the responses. The main focus of our statistical analysis is to assess the psychological stress based on the CPDI scores and the physician-specific distress score of the user responses to the questionnaires. Descriptive statistics such as mean, standard deviation, percentages, frequencies, T-test, and ANOVA-test were used for representing the insights of our analysis.

To assess the CPDI scores and psychological stress questionnaire scores, we divided the dataset into several categories and performed a T-test for each of them. For example, to understand the influence of gender on the CPDI score, we divided the dataset into two categories: Male and Female. After that, a T-test was performed to determine whether there are any significant differences among the responses to the CPDI score questions, including the overall CPDI scores of the male and female participants. A T-test was also performed for finding out the influence of gender on the psychological stress questionnaire. Similarly, the datasets were divided based on the following properties of the participants:

- District: Dhaka vs Other District
- Department: COVID-19 dedicated facility, ICU vs other
- Mode of Transport
- Experience in COVID-19 dedicated facility: Yes vs No
- Pre-existing Illness: Yes vs No
- Concomitant psychiatric disorders: Yes vs No
- Workplace: Tertiary level healthcare facility vs Other hospitals

For all these categories, T-tests were performed to find out any significant differences in the responses of the CPDI score and the responses of the psychological stress questionnaire. In all the T-tests, P-value of less than or equal to 0.05 was considered as the indicator of significant difference. The results of those T-tests helped us find valuable insights about the characteristics that influence the physicians’ distress level and psychological well-being. Apart from the T-test, mean values, standard deviations were also calculated for the CPDI scores of psychological stress questionnaires.

## Results

### Demographics Information

In our study, 237 doctors participated. Among them, 128 (53.78%) identified themselves as male participants, and 109 (45.79%) identified themselves as female participants. The average age of male participants is 30.10 with a standard deviation of ±5.23, and female participants have an average age of 29.03 with a standard deviation of ±3.33. Among the male participants, 39.1% reside in the capital city Dhaka, and 60.9% reside outside Dhaka. For female participants, 52.3% were found to be from Dhaka, while the rest were from outside of Dhaka. Based on the demography of Bangladesh, most of the female population with job prefers to stay close to their home [23] and our study also supports that actual scenario.

### COVID-19 Peritraumatic Distress Index of Physicians in Bangladesh

Our study shows, more than 78% respondents are suffering from peritraumatic psychological distress. COVID-19 Peritraumatic Distress Index (CPDI) scores of all participants show 50.82% of all participants are suffering moderate peritraumatic distress and 27.27% participants are suffering severe peritraumatic distress. Only 21.9% of the physicians were able to cope up with the pandemic; hence their peritraumatic stress identified as mild. Our study also indicates that female physicians showed significantly higher psychological distress compared to their male counterparts; 86.24% of female physicians are suffering from mild to severe psychological distress whereas 73.44% of male physicians are suffering from mild to severe psychological distress.

### Physician Specific Questionnaire

Physicians are constantly exposed to critical situations while treating COVID-19 patients. To better capture the physician-specific distress score, we used nine physician-specific COVID-19 psychological distress questionnaire from Wu et al. [4]. The questions are divided into three dimensions: (a) Risk awareness, which reflects the self-evaluation of the risk stoicism of the subject’s environment; (b) Physical and mental response, which reveals the stress response to the subject’s current environment; (c) Optimistic hope, which mirrors how much the subjects were confident in defeating the epidemic and the optimism of the current outbreak [4]. We studied how the stress score varies in different groups like genders, physicians with pre-existing illness, location, and prior job experience. **Table 1** summarizes the questions for specific group with corresponding scores which has p-value ≤ 0.05. We discuss the outcomes below:

**Table 1.**
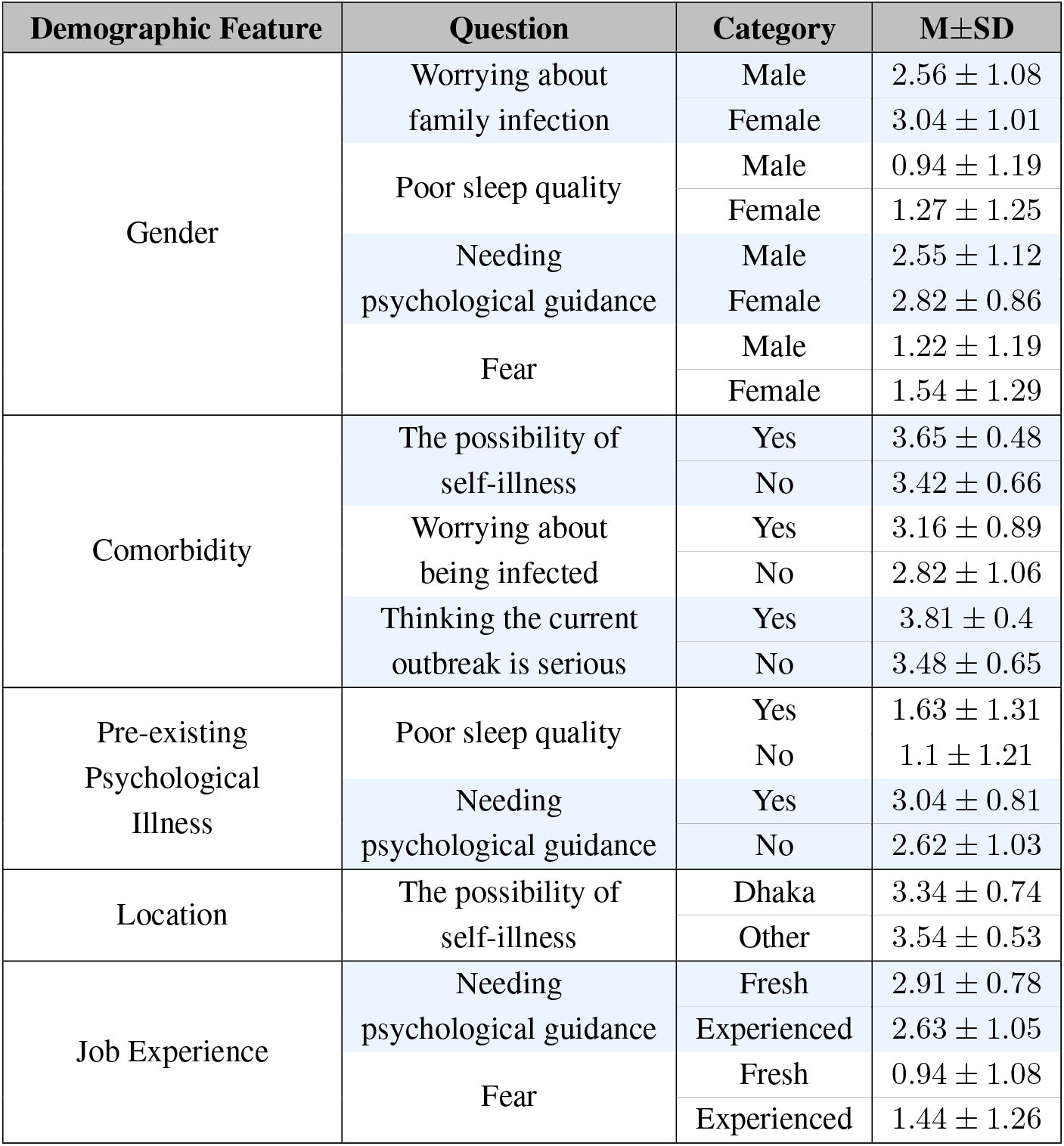
Summary of the physician specific questionnaire analysis based on the demographic features (p-value ≤ 0.05)

- **Physician specific stress index varies among the male and female doctors**. Our study shows female physicians suffer from more stress compared to their male counterparts. Most of the female physicians work near their family residence. Therefore, they are more worried about getting their family members infected by catching the infection for themselves. Female physicians also suffer from poor sleep quality and are more frightened of the outbreak compared to male physicians. They also feel that physiological guidance can be helpful to reduce the added stress.
- **Peritraumatic stress of the physicians with the pre-existing illness or comorbidity**. The higher fatality rate of COVID-19 has been seen for the patients older than 60 years of age or with preexisting illness. Our study shows that physicians with comorbidity suffer from significantly higher peritraumatic stress compared to young and healthy physicians. They are constantly worried about getting self-infection and believe that the outbreak may leave serious consequences on their health. Physicians with the pre-existing psychological illness also have a higher stress index. They have shown noteworthy sleep disorder issues during this outbreak and are in need of psychological guidance.
- **CPDI score varies among the physicians serving in urban and rural area**. Although the majority of the COVID-19 infection are in Dhaka and Chittagong city, physicians outside of Dhaka are more scared of getting self-infection. Our further inquiry has revealed that physicians outside of Dhaka don’t get access to enough protective equipment and don’t have access to advanced care unit facilities. Moreover, the patients outside Dhaka do not express their current condition clearly due to social stigma for which treating them as common patients increase the risk of silent infection [24]. Therefore, physicians outside the city areas think they have a higher chance of getting infected; and if they get infected then they might not receive proper treatment or ICU facilities unless they are transferred to Dhaka.

## Discussion and Recommendation

The major concern of the healthcare workers is to get access to personal protective equipment (PPE) while on duty. Many physicians do not have access to PPE, and even when they do, some of the PPE are below standard. Thus we urge the authorities to strictly maintain the PPE standard and to ensure the distribution of PPEs to all the frontline healthcare workers. Another major concern of the physicians in Bangladesh is that general population is not taking the pandemic situation seriously. **Figure 1(a)** shows that the drop in mobility of the population became negative, meaning the movement within the country has already become higher than the baseline. Thus proper steps and campaign to increase the awareness regarding the use of face mask, maintaining social distance and isolation by the authorities is necessary. Thus awareness about COVID-19 infection among the mass would help in the reduction of asymptomatic COVID-19 infection. This will certainly increase the trust and respect between patients and healthcare workers, which will potentially impact the frontliners’ CPDI score. We also forward three more policy recommendations to manage the healthcare workers’ CPDI score and thus helping them to manage both of their physical and mental stress.

### Evaluate Healthcare Workers’ CPDI and Comorbidities to Reduce Fatality Rate

On June 18th, the President of the Bangladesh Medical Association (BMA) mentioned that the fatality rate of Physicians in Bangladesh was 4% which is highest compared to any other country [6]. As of today (20th December), 119 doctors have lost their lives to COVID-19. Our study has revealed it’s not just the scarcity of the resources like PPEs or treatment facilities, poor management is also responsible for these higher fatality rates.

Our results show that about 20% physicians suffer from comorbidity or pre-existing illness. Therefore, if they get infected by COVID-19, the chances of becoming critically ill are very high which can result in deaths. Moreover, the added stress can put their physical and psychological health in jeopardy. We recommend evaluating the CPDI of each healthcare worker including physicians and nurses and take special care of those with severe CPDI scores and critical comorbidity issues. This way, the government of Bangladesh can ensure the healthcare facilities for COVID-19 frontline service providers.

### Reducing the Workload of Healthcare Workers with Higher CPDI

Bangladesh is a densely populated country. Even prior to the pandemic the country was suffering from a shortage of health-care workers per capita. According to WHO, there are an estimated 3.05 physicians and 1.07 nurses per 10,000 population in Bangladesh [12]. The COVID-19 pandemic had an overwhelming impact on the healthcare system of Bangladesh. Initially, very few hospitals had dedicated COVID-19 facilities and the testing capacity was also scarce. All the untested patients would rally in the nearest healthcare institutions. Therefore, the number of patients increased dramatically very quickly. All the vacation and holidays were canceled for the frontline workers, i.e. healthcare workers and police officers [7].

According to the survey participants of peritraumatic distress, the average working hours per month for the male participants is ∼ 150, while for the female participants it is ∼ 140. Though it may seem less number of working hours per month, but their duty schedule is arranged such a way that they do not get ample breaks between the duty hours for consecutive days. In some major COVID-19 dedicated hospitals physicians and nurses needs to work 15 consecutive days and followed by 15 days of self-isolation/quarantine. Thus, we recommend to identify those healthcare workers with severe CPDI score and reduce their workload in the frontline. We further suggest allowing those with severe CPDI to let them take specific hours of break every day to manage their physical and mental stress level. We speculate, such an intermittent break would help to reduce their current CPDI score significantly.

### Improving the Workplace Security and Dormitory Standard of Healthcare Workers

Bangladesh is a developing country. Workplace security and dormitory standard are still very low for the female workforce. From our study, we found that only 29.36% female physicians live outside their family residence while 47.66% male physicians live outside their family residence. The main reason behind this sheer difference is the increase in gender-based violence for female workers [25]. Thus, female physicians don’t feel comfortable living outside the family residence as the male population does.

Our study reveals that during the COVID-19 pandemic, physicians who are living with their family have a significantly higher stress index compared to those who live alone or live in a dormitory/hotel. Thus we recommend that authorities should take proper action to ensure that the temporary residence of female physicians maintain proper living standard with security and provide female-friendly facilities. In addition to these, we want to mention that female physicians with kids need daycare facilities for their children while living in temporary residence. Thus we suggest providing daycare facilities to the eligible kids for the physicians on duty.

## Concluding Remarks

Most of the countries of the world are overwhelmed by the COVID-19 pandemic. It has been an ultimate test of management for any governance, especially in the healthcare system. Bangladesh is not an exception in this manner. Even as a developing country with very limited resources Bangladesh is fighting the pandemic and the fatality rate has been low than the previous anticipation. Physicians, nurses, and other healthcare workers are giving their full effort to help people which sometimes results in losing their own lives. This sheer dedication should be used in a productive manner. In this paper, we analyzed the psychological stress of the physicians of Bangladesh during the pandemic based on the received feedback from COVID-19 designated physicians. Our study shows a huge percentage of the physicians are suffering from moderate to severe peritraumatic stress. If the situation doesn’t improve it may leave a crippling effect on the healthcare system. We also recommended some very important and yet easy to implement policies to reduce the peritraumatic stress of the physicians of Bangladesh. These policy recommendations were a result of the survey analysis and the suggestion from the COVID-19 designated physicians. We believe a proper implementation of these recommended policies can help significantly reduce the psychological stress of the healthcare workers of Bangladesh.

## Data Availability

The dataset will be made available on request.

## Data Availability Statement

The dataset will be made available on request.

## Ethics Statement

An informed consent of the study was taken from the participants. The criteria of selecting the participants was that only those adults who were directly involved to provide medical treatment physically to the COVID-19 patients in Bangladesh. To start filling out the questionnaire, participants were needed to choose “agree” option; otherwise, the questionnaire could not be filled out. Confidentiality was managed by placing anonymous coding of one self-report questionnaires. The study has been approved by the ethical clearance Committee of Sir Salimullah Medical College, Dhaka under the supervision of Md. Daharul Islam as the Principal Investigator of the project.

## Acknowledgments

We acknowledge the contribution of Dr. Enamul Hoque, Assistant Professor from Department of Physics, Shahjalal University of Science and Technology, Bangladesh; and, Dr. Shariful islam, Assistnat Professor from Department of Physics, North South University, Bangladesh especially for providing the COVID-19 daily cases data and SEIR modeling.

## References

1. Islam MS, Hoque ME, Amin MR. Integration of Kalman filter in the epidemiological model: a robust approach to predict COVID-19 outbreak in Bangladesh. medRxiv. 2020;.

2. Hoque E, Islam MS, Das SK, Mitra DK, Amin MR. Adjusted dynamics of COVID-19 pandemic due to herd immunity in Bangladesh. 2020;.

3. Qiu J, Shen B, Zhao M, Wang Z, Xie B, Xu Y. A nationwide survey of psychological distress among Chinese people in the COVID-19 epidemic: implications and policy recommendations. General Psychiatry. 2020 Mar;33(2):e100213. Available from: https://doi.org/10.1136/gpsych-2020-100213.

4. Wu W, Zhang Y, Wang P, Zhang L, Wang G, Lei G, et al. Psychological stress of medical staffs during outbreak of COVID-19 and adjustment strategy. Journal of Medical Virology. 2020 Jun;92(10):1962–1970. Available from: https://doi.org/10.1002/jmv.25914.

5. . Medi Voice [Accessed January 10, 2021]. 2020 Jun;Available from: https://medivoicebd.com/article/16689.

6. Coronavirus: Doctors’ mortality rate in Bangladesh ‘highest in the world’. United News of Bangladesh (UNB) [Accessed August 31, 2020]. 2020 Jun;Available from: https://www.msn.com/en-xl/news/other/coronavirus-doctors-mortality-rate-in-bangladesh-highest-in-the-world/ar-BB15M6KD.

7. All holidays of police, health and foreign ministry cancelled. Daily Bangladesh [Accessed December 25, 2020]. 2020 Mar;Available from: https://www.daily-bangladesh.com/english/All-holidays-of-police-health-and-foreign-ministry-cancelled/38990.

8. Mohiuddin AK. Diabetes Fact: Bangladesh Perspective. 2019 02;.

9. Andaleeb SS, Siddiqui N, Khandakar S. Patient satisfaction with health services in Bangladesh. Health Policy and Planning. 2007 May;22(4):263–273. Available from: https://doi.org/10.1093/heapol/czm017.

10. Al-Zaman MS. Healthcare Crisis in Bangladesh during the COVID-19 Pandemic. The American Journal of Tropical Medicine and Hygiene. 2020 Oct;103(4):1357–1359. Available from: https://doi.org/10.4269/ajtmh.20-0826.

11. Reza HM, Sultana F, Khan IO. Disruption of Healthcare Amid COVID-19 Pandemic in Bangladesh. The Open Public Health Journal. 2020 Sep;13(1):438–440. Available from: https://doi.org/10.2174/1874944502013010438.

12. Bangladesh. World Health Organization [Accessed December 17, 2020]. 2012 Apr;Available from: https://www.who.int/workforcealliance/countries/bgd/en/.

13. Maunder R, Lancee W, Balderson K, Bennett J, Borgundvaag B, Evans S, et al. Long-term Psychological and Occupational Effects of Providing Hospital Healthcare during SARS Outbreak. Emerging Infectious Diseases. 2006;12(12):1924–1932. Available from: https://doi.org/10.3201/eid1212.060584.

14. Dutheil F, Mondillon L, Navel V. PTSD as the second tsunami of the SARS-Cov-2 pandemic. Psychological Medicine. 2020;p. 1–2.

15. Benfante A, Tella MD, Romeo A, Castelli L. Traumatic Stress in Healthcare Workers During COVID-19 Pandemic: A Review of the Immediate Impact. Frontiers in Psychology. 2020 Oct;11. Available from: https://doi.org/10.3389/fpsyg.2020.569935.

16. Shanafelt TD, Boone S, Tan L, Dyrbye LN, Sotile W, Satele D, et al. Burnout and Satisfaction With Work-Life Balance Among US Physicians Relative to the General US Population. Archives of Internal Medicine. 2012 Oct;172(18):1377. Available from: https://doi.org/10.1001/archinternmed.2012.3199.

17. Shanafelt TD, Hasan O, Dyrbye LN, Sinsky C, Satele D, Sloan J, et al. Changes in Burnout and Satisfaction With Work-Life Balance in Physicians and the General US Working Population Between 2011 and 2014. Mayo Clinic Proceedings. 2015 Dec;90(12):1600–1613. Available from: https://doi.org/10.1016/j.mayocp.2015.08.023.

18. Rotenstein LS, Ramos MA, Torre M, Segal JB, Peluso MJ, Guille C, et al. Prevalence of Depression, Depressive Symptoms, and Suicidal Ideation Among Medical Students. JAMA. 2016 Dec;316(21):2214. Available from: https://doi.org/10.1001/jama.2016.17324.

19. Mata DA, Ramos MA, Bansal N, Khan R, Guille C, Angelantonio ED, et al. Prevalence of Depression and Depressive Symptoms Among Resident Physicians. JAMA. 2015 Dec;314(22):2373. Available from: https://doi.org/10.1001/jama.2015.15845.

20. Coronavirus Cases. Worldometer [Accessed December 16, 2020]. 2020 December;Available from: https://www.worldometers.info/coronavirus/.

21. The Challanges of Good Governance to Combat COVID-19. Transparency International Bangladesh [Accessed December 15, 2020]. 2020 June;Available from: https://www.ti-bangladesh.org/beta3/images/2020/report/covid-19/Covid-Resp-Track-Full-BN-15062020.pdf.

22. 79% of patients get treatment over the phone. Prothomalo [Accessed September 22, 2020]. 2020 May;Available from: https://www.prothomalo.com/bangladesh/article/1653078/.

23. Kibria N. CULTURE, SOCIAL CLASS, AND INCOME CONTROL IN THE LIVES OF WOMEN GARMENT WORKERS IN BANGLADESH. Gender & Society. 1995 Jun;9(3):289–309. Available from: https://doi.org/10.1177/089124395009003003.

24. Mahmud A, Islam MR. Social Stigma as a Barrier to Covid-19 Responses to Community Well-Being in Bangladesh. International Journal of Community Well-Being. 2020 Aug;Available from: https://doi.org/10.1007/s42413-020-00071-w.

25. Akhter Z. EVE TEASING, TEARS OF THE GIRLS: Bangladesh Open University towards Women Empowerment; 2013..

